# Heterogeneous association between maternal haemoglobin concentration and child haemoglobin levels: a quantile regression analysis using multi-country nationally representative data

**DOI:** 10.64898/2026.11.02.26367046

**Authors:** Dickson A Amugsi, Daniel Osuka, Bonventure Mwangi, Sofia Jomo, Stephen Ekiru, Francisca Achaliwie, Calistus Wilunda

## Abstract

**Background:** Low haemoglobin concentration, or anaemia, is a major global public health issue, particularly severe in sub-Saharan Africa, including East Africa, where prevalence rates are alarmingly high. This situation requires a comprehensive analysis to identify the key factors contributing to this high prevalence, especially in East Africa. This study uses robust quantile regression to examine the relationship between maternal haemoglobin concentration and child haemoglobin levels in three East African countries.

**Methods:** The study analysed data from the Demographic and Health Surveys of three East African countries: Ethiopia, Tanzania, and Uganda. These data are nationally representative and are collected every five years from low- and middle-income countries. The analysis included a total sample of 6,883 children and their mothers. The primary outcome variable was the child’s haemoglobin concentration. We used quantile regression to model the effects of maternal haemoglobin concentration on child haemoglobin levels, adjusting for potential confounders.

**Results:** The results showed that a one-unit increase in maternal haemoglobin (Hb) concentration was associated with a unit increase in child haemoglobin concentration as follows: In Tanzania, the increases were 0.19, 0.20, 0.19, and 0.17 at the 5th, 10th, 25th, and 50th quantiles, respectively. In Ethiopia, the increases were 0.23, 0.22, 0.22, and 0.17 at the same quantiles. In Uganda, the increases were 0.19, 0.19, 0.18, and 0.19 at these quantiles. In contrast, a one-unit increase in maternal Hb concentration was associated with a smaller increase in child Hb concentration, ranging from 0.11 to 0.13 at the upper quantiles (75th and 90th) across all three countries. Overall, the most significant effects of maternal Hb concentration on child Hb levels were observed at the lower end of the conditional Hb distribution in each of the three countries.

**Conclusion:** Maternal haemoglobin (Hb) concentration is positively associated with children’s haemoglobin levels across all countries studied. The strongest association is among children in the lower quantiles, who are at higher risk of anaemia. In contrast, the association is less pronounced among those in the upper quantiles. Therefore, improving maternal Hb is likely to benefit children in the lower quantiles more than those in the upper quantiles. Programmes aimed at addressing child anaemia should consider these differing effects of maternal Hb concentration.

## Background

Childhood anaemia, commonly known as low haemoglobin (Hb) concentration, reflects a reduced red blood cell count or diminished oxygen-carrying capacity (1). This condition is a major public health problem, affecting an estimated 40% of children aged 6 to 59 months worldwide (1). The situation is particularly severe in sub-Saharan Africa, where the prevalence is estimated at 64%, and in some countries, it exceeds 80% (2, 3). In East Africa, the average prevalence is 55.6% (4). In some specific East African countries, such as Tanzania, Ethiopia, and Uganda, the burden is 56%, 52%, and 52%, respectively (5). This substantial prevalence underscores the urgent need to identify the primary determinants of childhood anaemia in the region, so that effective interventions can be designed and targeted appropriately.

Low Hb concentration can have significant short- and long-term effects on children’s health. Anaemia negatively affects various aspects of a child’s wellbeing, including developmental delays, impaired cognitive development (which can reduce learning ability and school performance), weakened immunity, fatigue, difficulty concentrating, lethargy, increased mortality, and heightened susceptibility to infections (1). Moreover, childhood anaemia is linked to a reduced ability to fight infections, contributing to morbidity and mortality among children (1).

The extant literature suggests that various factors, including deficiencies in essential nutrients such as iron, folate, vitamin B12, and copper, influence low haemoglobin levels in children. (6). Other contributing elements include insufficient dietary intake, poor nutrient absorption, and increased physiological needs during periods of rapid growth, along with key sociodemographic factors at the maternal and household levels (7). However, this study focuses on the effects of maternal haemoglobin concentration on their children’s haemoglobin levels. This focus is justified by evidence of a strong relationship between maternal and child haemoglobin concentrations (7, 8).

Several studies have also documented a strong association between maternal haemoglobin concentration and childhood anaemia (low haemoglobin concentration) (3, 7–12). Compared to children of non-anaemic mothers, children of anaemic mothers are approximately 13.5% more likely to be anaemic themselves (9). A cross-sectional study conducted in rural Zimbabwe found a significant link between maternal anaemia and anaemia in children aged 6 to 59 months, indicating a cross-generational relationship (8). The research emphasised that improving maternal nutritional status is essential for reducing the high rates of child anaemia in low-income settings (8). This intergenerational connection largely stems from shared nutritional deficiencies, socioeconomic factors, and infections, which often lower haemoglobin levels (9). Many other studies have identified maternal anaemia as a key driver of child haemoglobin concentration levels. In a study using data from 47 Sub-Saharan African (SSA) countries, researchers found that the highest anaemia prevalence occurred in children whose mothers were moderately or severely anaemic, at 76.8% and 76.7%, respectively (3). This indicates that children of anaemic mothers are more likely to experience higher levels of anaemia compared to children whose mothers are not anaemic.

The existing literature on the relationship between maternal and child anaemia levels abounds. However, the analytical methods used in previous studies have significant limitations. Most of these studies have relied on Ordinary Least Squares (OLS) or logistic regression to estimate the effects of maternal Hb concentration on child Hb levels. These approaches may overlook the varying effects that maternal Hb concentration can have on child Hb levels. This variability highlights the need for a more comprehensive analysis to understand how maternal Hb concentration influences child Hb status across the Hb concentration distribution. Therefore, this study employs quantile regression, a method that allows for the examination of maternal Hb effects at various points within the conditional distribution of child Hb concentration. Such an analysis is currently lacking in the field of child anaemia research.

## Materials and Methods

### Data sources, study design and population

This analysis utilised the Demographic and Health Surveys (DHS) data sets (13) from three East African countries: Tanzania, Ethiopia, and Uganda. We chose these countries because they are the countries in EA which have data on child and maternal Haemoglobin. The DHS survey design is consistent across these countries, allowing meaningful comparisons. The DHS employed a two-stage sampling design. In the first stage, clusters were selected from an updated master sampling frame based on the National Population and Housing Census of each country. Clusters were chosen using systematic sampling with probability proportional to size. Following this, household listings were conducted in all selected clusters to create a sampling frame for the second stage. In the second stage, households from the listed households in each cluster were systematically sampled, with selections made randomly from the list. This approach ensures an adequate number of completed individual interviews, allowing reliable estimates of key indicators with acceptable precision. All women aged 15 to 49, along with their children aged 0-59 months, in the selected households were eligible to participate in the surveys as long as they were either usual residents of the household or visitors present in the household on the night before the survey. This analysis focuses on children aged 6 to 59 months, the eligible age for inclusion in the Hb data collection. The total sample sizes used in this analysis were 1797, 3486, and 1600 mother-child dyads, representing Tanzania, Ethiopia, and Uganda, respectively.

### Ethics

The DHS study was conducted in accordance with international ethical best practices (14). Thus, data collectors were trained to respect the rights of individuals participating in the research. During data collection, participants were informed of their right to decide whether to take part in the study. They were also told they could abstain or withdraw (e.g., discontinue an interview) at any time without repercussions. The study’s risks and benefits were clearly explained, along with measures taken to mitigate potential risks. Well-trained investigators conducted the surveys using an approved protocol. The relevant national Ethical Review Committees in each country, as well as the Institutional Review Board of ICF International in the USA, reviewed and approved the study protocols, including those for biomarker collection.

The DHS research team obtained written informed consent from mothers before the survey, and for children aged 6–59 months, from their parents or guardians. The team shared biomarker results with participants. For example, the Hb test results for all children tested in the household were recorded in a malaria and anaemia brochure and given to the parent or responsible adult. Additionally, an anaemia referral form was given to facilitate immediate treatment at a nearby health facility for children with an Hb level below 7.0 g/dl. The authors requested and received permission from the DHS programme to use the data. Because the data were fully anonymous, the authors did not need to seek additional ethical clearance.

## Measures

### Outcome variable(s)

The outcome variable was child Hb concentration. The DHS collected blood specimens from all children aged 6 to 59 months for anaemia testing. A drop of blood was obtained through a finger prick for older children and a heel prick for those aged 6 to 11 months. The blood was collected in a microcuvette. Hb analysis was performed on-site using a portable battery-powered HaemoCue analyser. The Hb data were categorised as follows: mild anaemia (Hb concentration of 10.0 to 10.9 grams per deciliter (g/dL)), moderate anaemia (Hb concentration of 7.0 to 9.9 g/dL), and severe anaemia (Hb concentration of less than 7.0 g/dL). In this analysis, Hb concentration was treated as a continuous variable due to the analytical strategy employed.

### Explanatory variables

#### Primary explanatory variable

The exposure was maternal Hb concentration (g/dL), a continuous variable. The DHS collected blood samples from non-pregnant mothers who consented to participate. Blood was obtained from a finger prick and analysed on-site using a portable HaemoCue analyser.

#### Covariates

Covariates were selected based on prior literature and organised into four levels. At the child level, the covariates included sex, age in months, and vitamin A supplementation during the six months preceding the survey. Vitamin A supplementation was specifically chosen as a covariate because it raises haemoglobin levels and lowers the risk of anaemia by 26% (15). At the maternal level, the covariates were years of education, age, body mass index (BMI), current employment status, breastfeeding status, and antenatal care (ANC) attendance. Maternal BMI was categorised as follows: underweight (<18.5 kg/m²), normal weight (18.5–24.9 kg/m²), overweight (25.0–29.9 kg/m²), or obese (≥30 kg/m²). We excluded extreme and implausible values before categorisation. We classified ANC attendance as adequate (four or more visits) or inadequate (fewer than four visits), in line with WHO recommendations in effect during each survey (16).

At the household level, the covariates included the household wealth index, the sex of the household head, household size, the primary decision-maker for large purchases, and the highest education level of the husband or partner. At the community level, we also considered place of residence (urban or rural).

### Statistical analysis

Analyses were performed in Stata version 17 using the svyset command to account for the complex survey design. We computed weighted descriptive statistics for the analytic sample in each country, reporting means and standard deviations for continuous variables and weighted proportions for categorical variables. To assess crude associations, we first ran bivariate OLS regressions for each covariate against child haemoglobin concentration. We then included all covariates with evidence of association in bivariate analysis or established theoretical relevance in a multivariable, survey-weighted OLS regression, estimated using *svy: reg*, which provides unbiased coefficient estimates and design-corrected standard errors under the complex sampling scheme. The multivariable OLS model estimated the mean difference in child haemoglobin concentration associated with a one-unit increase in maternal haemoglobin concentration, holding all other covariates constant. We assessed potential multicollinearity among covariates using variance inflation factors (VIF).

To test whether the association between maternal and child haemoglobin concentration varied across the conditional distribution of child haemoglobin, we estimated quantile regression (QR) models at the 5th, 10th, 25th, 50th, 75th, and 90th percentiles. Unlike OLS, which estimates effects at the conditional mean, QR characterises the relationship at multiple points of the outcome distribution, offering insight into whether the association is stronger among children with low haemoglobin—where the clinical implications are greatest—than among those with higher levels. The model was specified as:

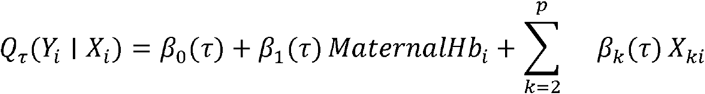

where *Q_τ_ (Y_i_ | X_i_)* is the *τ*-th conditional quantile of child haemoglobin concentration, β_1_(τ) is the effect of maternal haemoglobin at quantile τ, and *X_ki_* is the vector of covariates. We applied sampling weights in all QR models. We excluded marital status from the final models in all three countries because of limited variability, with over 83% of mothers reporting they were currently married or cohabiting.

All analyses accounted for the complex survey design of the DHS. We constructed sampling weights by dividing the DHS weight variable (v005) by 1,000,000. We declared the survey design in Stata using *svyset*, specifying primary sampling units (v021) and sampling strata (v022), so variance estimates reflected the stratified cluster structure across all descriptive and regression analyses.

## Results

### Descriptive statistics

Hb concentration among children in the three countries was similar, with Uganda showing a slightly higher average of 10.33 ± 1.79 g/dL. Women in Ethiopia had the highest Hb concentration (12.54 ± 2.59 g/dL), while women in Tanzania had the lowest (12.10±2.36). In terms of maternal education, women in Ethiopia had the fewest years of schooling, averaging 2.06 ± 5.03 years, compared with women in Tanzania (6.20 ± 4.56 years) and Uganda (6.25 ± 4.89 years). Additionally, women in Ethiopia had the lowest BMI (20.50 ± 5.14 kg/m²), while women in Tanzania and Uganda had higher averages of 23.22 ± 1.66 and 23.02 ± 5.49, respectively. Tanzania also had fewer female children (47.6%) than Ethiopia (50.5%) and Uganda (51.2%). Uganda had the highest percentage of children receiving Vitamin A (76%), while Ethiopia had the lowest (42%).

### Quantile regression analysis

Tables 2–4 present the results of the multivariable QR analysis for Tanzania, Ethiopia, and Uganda. To facilitate comparison with the QR findings, we have also included OLS estimates in all tables. The OLS analysis indicated a positive association between maternal haemoglobin (Hb) concentration and their children’s Hb levels across all three countries and quantiles. Thus, a unit increase in maternal Hb concentration corresponded to a one-unit increase in the child’s Hb level. However, the QR analysis revealed that the effects of maternal Hb concentration on child Hb levels varied across points in the conditional Hb distribution. In Tanzania, the strongest association between maternal Hb concentration and child Hb levels was observed in the lower four quantiles, while the weakest associations were noted among children in the upper two quantiles. A similar pattern emerged in analyses of data from Ethiopia and Uganda, where maternal Hb concentration was positively associated with child Hb levels across all quantiles. Again, the strongest association was found among children in the lower quantiles, and the weakest association among those in the upper two quantiles. Specifically, a one-unit increase in maternal Hb concentration was associated with the following increases in child Hb concentration: In Tanzania, 0.19, 0.20, 0.19, and 0.17 at the 5th, 10th, 25th, and 50th quantiles, respectively. In Ethiopia, the increases were 0.23, 0.22, 0.22, and 0.17 at the same quantiles. In Uganda, the increases were 0.19, 0.19, 0.18, and 0.19 at the respective quantiles. In contrast, a one-unit increase in maternal Hb concentration was associated with a smaller increase in child Hb concentration, ranging from 0.11 to 0.13 at the upper quantiles (75th and 90th) across all three countries. Overall, the most significant effects of maternal Hb concentration on child Hb levels were observed at the lower end of the conditional Hb distribution in each of the three countries.

**Table 1:** Descriptive results of socio-demographic factors.

|  | **Tanzania (n=1797)** | **Ethiopia (n=3486)** | **Uganda (n=1600)** |
|---|---|---|---|
| **Variable** | **Mean (SD) or n (%)** | **Mean (SD) or n (%)** | **Mean (SD) or n (%)** |
| Child Hb (g/dL) | 10.18 (1.86) | 10.15 (2.38) | 10.33(1.79) |
| Maternal Hb (g/dL) | 12.10 (2.36) | 12.54 (2.59) | 12.43(1.98) |
| Child Age (months) | 19.00 (10.24) | 18.71 (12.14) | 18.71(9.93) |
| Maternal Education (years) | 6.20 (4.86) | 2.06 (5.03) | 6.25(4.89) |
| Maternal Age (years) | 29.11 (8.39) | 29.08 (9.44) | 28.39(7.52) |
| Maternal BMI (kg/m<sup>2</sup>) | 23.22 (1.66) | 20.50 (5.14) | 23.02(5.49) |
| Child Sex |  |  |  |
| Female | 856 (47.6%) | 1,761 (50.5%) | 821 (51.2%) |
| Male | 942 (52.4%) | 1,728 (49.5%) | 781 (48.8%) |
| Received Vitamin A (last 6 months) |  |  |  |
| Yes | 1120 (62.3%) | 1,463 (42.0%) | 1,217 (76.0%) |
| No | 678 (37.7%) | 2,026 (58.0%) | 385 (24.0%) |
| Wealth Index |  |  |  |
| Poorest | 392 (21.8%) | 797 (22.8%) | 353 (22.0%) |
| Poorer | 347 (19.3%) | 777 (22.3%) | 342 (21.3%) |
| Middle | 327 (18.2%) | 743 (21.3%) | 306 (19.1%) |
| Richer | 419 (23.3%) | 642 (18.4%) | 284 (17.7%) |
| Richest | 313 (17.4%) | 530 (15.2%) | 317 (19.8%) |

**Table 2.**
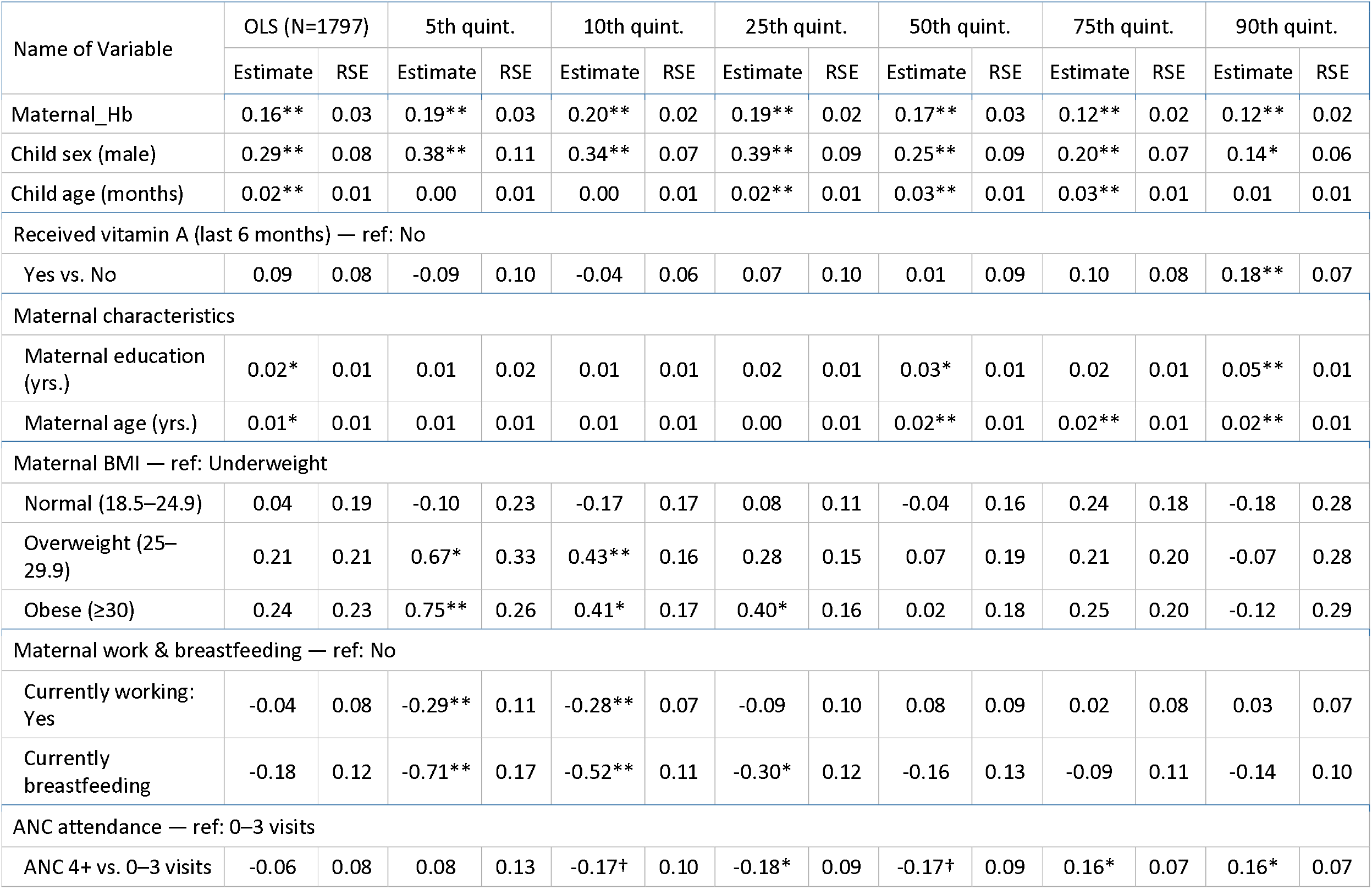
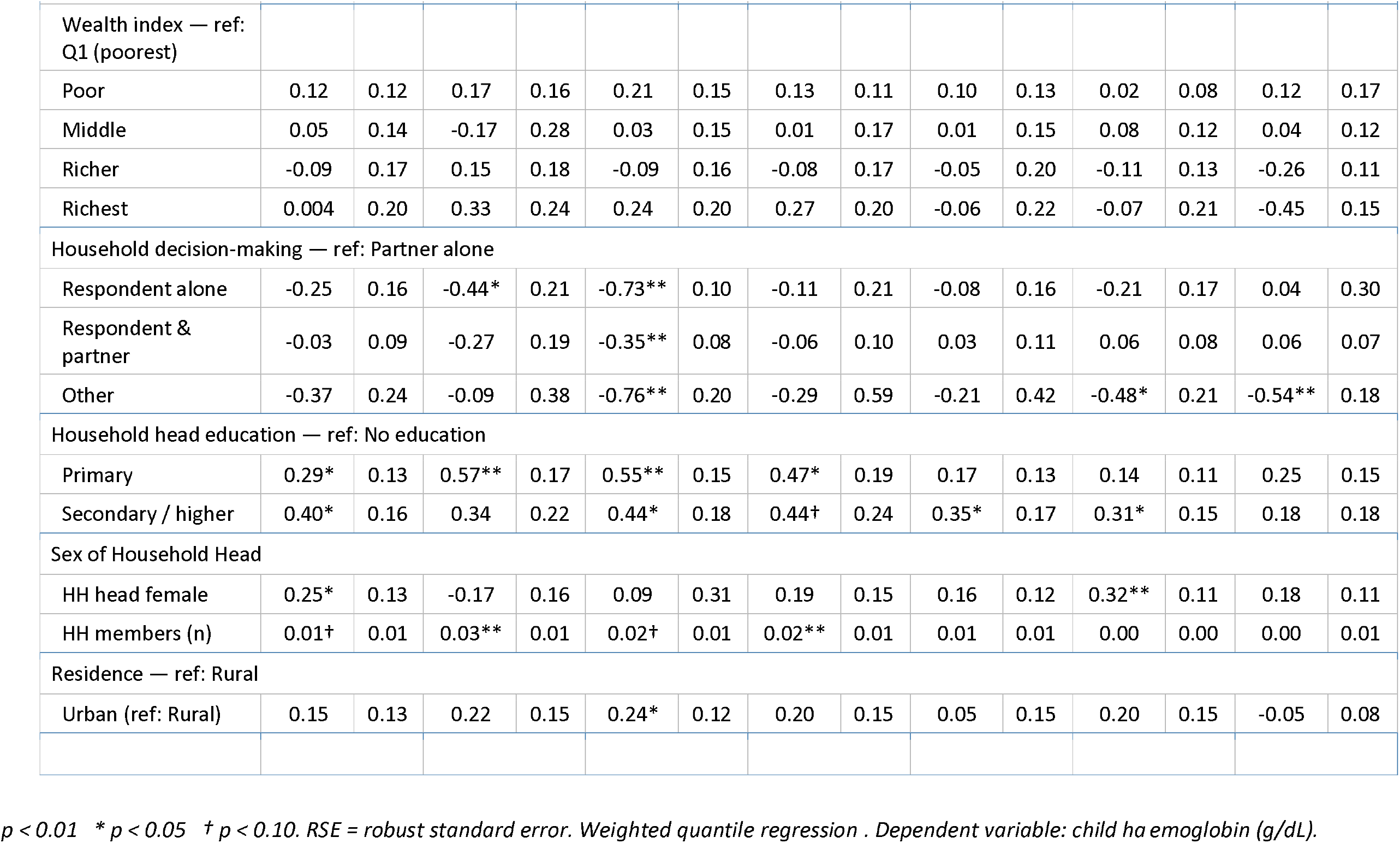
Multivariable analysis of the association between maternal and child haemoglobin concentration, Tanzania (N = 1797).

| Name of Variable | OLS (N=1797) |  | 5th quint. |  | 10th quint. |  | 25th quint. |  | 50th quint. |  | 75th quint. |  | 90th quint. |  |
|---|---|---|---|---|---|---|---|---|---|---|---|---|---|---|
|  | Estimate | RSE | Estimate | RSE | Estimate | RSE | Estimate | RSE | Estimate | RSE | Estimate | RSE | Estimate | RSE |
| Maternal_Hb | 0.16** | 0.03 | 0.19** | 0.03 | 0.20** | 0.02 | 0.19** | 0.02 | 0.17** | 0.03 | 0.12** | 0.02 | 0.12** | 0.02 |
| Child sex (male) | 0.29** | 0.08 | 0.38** | 0.11 | 0.34** | 0.07 | 0.39** | 0.09 | 0.25** | 0.09 | 0.20** | 0.07 | 0.14* | 0.06 |
| Child age (months) | 0.02** | 0.01 | 0.00 | 0.01 | 0.00 | 0.01 | 0.02** | 0.01 | 0.03** | 0.01 | 0.03** | 0.01 | 0.01 | 0.01 |
| Received vitamin A (last 6 months) — ref: No |  |  |  |  |  |  |  |  |  |  |  |  |  |  |
| Yes vs. No | 0.09 | 0.08 | -0.09 | 0.10 | -0.04 | 0.06 | 0.07 | 0.10 | 0.01 | 0.09 | 0.10 | 0.08 | 0.18** | 0.07 |
| Maternal characteristics |  |  |  |  |  |  |  |  |  |  |  |  |  |  |
| Maternal education (yrs.) | 0.02* | 0.01 | 0.01 | 0.02 | 0.01 | 0.01 | 0.02 | 0.01 | 0.03* | 0.01 | 0.02 | 0.01 | 0.05** | 0.01 |
| Maternal age (yrs.) | 0.01* | 0.01 | 0.01 | 0.01 | 0.01 | 0.01 | 0.00 | 0.01 | 0.02** | 0.01 | 0.02** | 0.01 | 0.02** | 0.01 |
| Maternal BMI — ref: Underweight |  |  |  |  |  |  |  |  |  |  |  |  |  |  |
| Normal (18.5–24.9) | 0.04 | 0.19 | -0.10 | 0.23 | -0.17 | 0.17 | 0.08 | 0.11 | -0.04 | 0.16 | 0.24 | 0.18 | -0.18 | 0.28 |
| Overweight (25–29.9) | 0.21 | 0.21 | 0.67* | 0.33 | 0.43** | 0.16 | 0.28 | 0.15 | 0.07 | 0.19 | 0.21 | 0.20 | -0.07 | 0.28 |
| Obese (≥30) | 0.24 | 0.23 | 0.75** | 0.26 | 0.41* | 0.17 | 0.40* | 0.16 | 0.02 | 0.18 | 0.25 | 0.20 | -0.12 | 0.29 |
| Maternal work & breastfeeding — ref: No |  |  |  |  |  |  |  |  |  |  |  |  |  |  |
| Currently working: Yes | -0.04 | 0.08 | -0.29** | 0.11 | -0.28** | 0.07 | -0.09 | 0.10 | 0.08 | 0.09 | 0.02 | 0.08 | 0.03 | 0.07 |
| Currently breastfeeding | -0.18 | 0.12 | -0.71** | 0.17 | -0.52** | 0.11 | -0.30* | 0.12 | -0.16 | 0.13 | -0.09 | 0.11 | -0.14 | 0.10 |
| ANC attendance — ref: 0–3 visits |  |  |  |  |  |  |  |  |  |  |  |  |  |  |
| ANC 4+ vs. 0–3 visits | -0.06 | 0.08 | 0.08 | 0.13 | -0.17† | 0.10 | -0.18* | 0.09 | -0.17† | 0.09 | 0.16* | 0.07 | 0.16* | 0.07 |
| Wealth index — ref: Q1 (poorest) |  |  |  |  |  |  |  |  |  |  |  |  |  |  |
| Poor | 0.12 | 0.12 | 0.17 | 0.16 | 0.21 | 0.15 | 0.13 | 0.11 | 0.10 | 0.13 | 0.02 | 0.08 | 0.12 | 0.17 |
| Middle | 0.05 | 0.14 | -0.17 | 0.28 | 0.03 | 0.15 | 0.01 | 0.17 | 0.01 | 0.15 | 0.08 | 0.12 | 0.04 | 0.12 |
| Richer | -0.09 | 0.17 | 0.15 | 0.18 | -0.09 | 0.16 | -0.08 | 0.17 | -0.05 | 0.20 | -0.11 | 0.13 | -0.26 | 0.11 |
| Richest | 0.004 | 0.20 | 0.33 | 0.24 | 0.24 | 0.20 | 0.27 | 0.20 | -0.06 | 0.22 | -0.07 | 0.21 | -0.45 | 0.15 |
| Household decision-making — ref: Partner alone |  |  |  |  |  |  |  |  |  |  |  |  |  |  |
| Respondent alone | -0.25 | 0.16 | -0.44* | 0.21 | -0.73** | 0.10 | -0.11 | 0.21 | -0.08 | 0.16 | -0.21 | 0.17 | 0.04 | 0.30 |
| Respondent & partner | -0.03 | 0.09 | -0.27 | 0.19 | -0.35** | 0.08 | -0.06 | 0.10 | 0.03 | 0.11 | 0.06 | 0.08 | 0.06 | 0.07 |
| Other | -0.37 | 0.24 | -0.09 | 0.38 | -0.76** | 0.20 | -0.29 | 0.59 | -0.21 | 0.42 | -0.48* | 0.21 | -0.54** | 0.18 |
| Household head education — ref: No education |  |  |  |  |  |  |  |  |  |  |  |  |  |  |
| Primary | 0.29* | 0.13 | 0.57** | 0.17 | 0.55** | 0.15 | 0.47* | 0.19 | 0.17 | 0.13 | 0.14 | 0.11 | 0.25 | 0.15 |
| Secondary / higher | 0.40* | 0.16 | 0.34 | 0.22 | 0.44* | 0.18 | 0.44† | 0.24 | 0.35* | 0.17 | 0.31* | 0.15 | 0.18 | 0.18 |
| Sex of Household Head |  |  |  |  |  |  |  |  |  |  |  |  |  |  |
| HH head female | 0.25* | 0.13 | -0.17 | 0.16 | 0.09 | 0.31 | 0.19 | 0.15 | 0.16 | 0.12 | 0.32** | 0.11 | 0.18 | 0.11 |
| HH members (n) | 0.01† | 0.01 | 0.03** | 0.01 | 0.02† | 0.01 | 0.02** | 0.01 | 0.01 | 0.01 | 0.00 | 0.00 | 0.00 | 0.01 |
| Residence — ref: Rural |  |  |  |  |  |  |  |  |  |  |  |  |  |  |
| Urban (ref: Rural) | 0.15 | 0.13 | 0.22 | 0.15 | 0.24* | 0.12 | 0.20 | 0.15 | 0.05 | 0.15 | 0.20 | 0.15 | -0.05 | 0.08 |

*p < 0.01 \* p < 0.05 <sup>†</sup> p < 0.10. RSE = robust standard error. Weighted quantile regression . Dependent variable: child ha emoglobin (g/dL).*

**Table 3.** Multivariable analysis of the association between maternal and child haemoglobin concentration Ethiopia (N = 3,486).

| Name of Variable | OLS |  | 5th quant. |  | 10th quant. |  | 25th quant. |  | 50th quant. |  | 75th quant. |  | 90th quant. |  |
|---|---|---|---|---|---|---|---|---|---|---|---|---|---|---|
|  | Estimate | RSE | Estimate | RSE | Estimate | RSE | Estimate | RSE | Estimate | RSE | Estimate | RSE | Estimate | RSE |
| Maternal_Hb | 0.17** | 0.03 | 0.23** | 0.02 | 0.22** | 0.04 | 0.22** | 0.03 | 0.17** | 0.03 | 0.13** | 0.02 | 0.13** | 0.02 |
| Child sex (male) | 0.11 | 0.08 | 0.31* | 0.15 | 0.38** | 0.11 | 0.31** | 0.10 | 0.01 | 0.09 | 0.02 | 0.08 | 0.00 | 0.06 |
| Child age (months) | 0.03** | 0.01 | 0.03** | 0.01 | 0.02** | 0.01 | 0.03** | 0.01 | 0.04** | 0.01 | 0.03** | 0.00 | 0.03** | 0.00 |
| Received vitamin A (last 6 months) — ref: No |  |  |  |  |  |  |  |  |  |  |  |  |  |  |
| Yes vs. No | -0.01 | 0.08 | -0.03 | 0.12 | 0.07 | 0.12 | -0.04 | 0.10 | -0.08 | 0.09 | 0.10 | 0.08 | 0.03 | 0.07 |
| Maternal characteristics |  |  |  |  |  |  |  |  |  |  |  |  |  |  |
| Maternal education (yrs.) | 0.00 | 0.01 | -0.02 | 0.03 | 0.00 | 0.02 | 0.00 | 0.02 | 0.01 | 0.01 | 0.00 | 0.01 | -0.01 | 0.01 |
| Maternal age (yrs.) | 0.01 | 0.01 | -0.01 | 0.01 | 0.00 | 0.01 | 0.01 | 0.01 | 0.01 | 0.01 | 0.02** | 0.01 | 0.02** | 0.01 |
| Maternal BMI — ref: Underweight |  |  |  |  |  |  |  |  |  |  |  |  |  |  |
| Normal (18.5–24.9) | 0.08 | 0.10 | 0.20† | 0.12 | 0.05 | 0.18 | 0.15 | 0.12 | 0.06 | 0.10 | -0.07 | 0.09 | 0.05 | 0.07 |
| Overweight (25–29.9) | 0.16 | 0.16 | 0.53* | 0.23 | 0.60† | 0.33 | 0.24 | 0.16 | -0.04 | 0.21 | -0.12 | 0.17 | 0.11 | 0.10 |
| Obese (≥30) | 0.29 | 0.20 | 1.10 | 1.52 | 0.87 | 0.85 | 0.80** | 0.24 | 0.33 | 0.56 | -0.20 | 0.19 | -0.46 | 1.44 |
| Maternal work & breastfeeding — ref: No |  |  |  |  |  |  |  |  |  |  |  |  |  |  |
| Currently working: Yes | 0.16† | 0.08 | 0.35** | 0.11 | 0.20† | 0.12 | 0.17 | 0.11 | 0.15 | 0.09 | 0.07 | 0.08 | 0.09 | 0.08 |
| Currently breastfeeding | 0.16 | 0.11 | -0.11 | 0.13 | 0.13 | 0.14 | 0.23 | 0.15 | 0.16 | 0.12 | 0.08 | 0.09 | 0.05 | 0.10 |
| ANC attendance — ref: 0–3 visits |  |  |  |  |  |  |  |  |  |  |  |  |  |  |
| ANC 4+ vs. 0–3 visits | 0.07 | 0.09 | 0.28 | 0.18 | 0.22 | 0.12 | 0.03 | 0.11 | 0.05 | 0.10 | -0.05 | 0.09 | 0.03 | 0.07 |
| Wealth index — ref: Poorest |  |  |  |  |  |  |  |  |  |  |  |  |  |  |
| Poor | 0.39** | 0.13 | 0.15 | 0.17 | 0.57* | 0.24 | 0.53** | 0.16 | 0.60** | 0.14 | 0.29** | 0.11 | 0.03 | 0.10 |
| Middle | 0.63** | 0.14 | 0.99** | 0.17 | 1.15** | 0.17 | 0.77** | 0.16 | 0.55** | 0.14 | 0.47** | 0.11 | 0.22† | 0.12 |
| Rich | 0.52** | 0.16 | 0.56** | 0.18 | 0.99** | 0.22 | 0.40* | 0.16 | 0.52** | 0.16 | 0.46** | 0.14 | 0.34** | 0.12 |
| Richest | 0.70** | 0.18 | 1.55** | 0.29 | 1.37** | 0.22 | 1.00** | 0.23 | 0.60** | 0.15 | 0.58** | 0.17 | 0.28* | 0.12 |
| Household decision-making — ref: Partner alone |  |  |  |  |  |  |  |  |  |  |  |  |  |  |
| Respondent alone | 0.11 | 0.16 | -0.03 | 0.26 | 0.02 | 0.18 | -0.03 | 0.22 | 0.20 | 0.18 | 0.07 | 0.10 | -0.08 | 0.16 |
| Respondent & partner | 0.18† | 0.10 | 0.19 | 0.17 | 0.33* | 0.13 | 0.14 | 0.14 | 0.15 | 0.11 | 0.23* | 0.10 | 0.20 | 0.15 |
| Other | 0.56 | 0.92 | 0.79 | 2.22 | 0.62 | 1.56 | 0.12 | 0.31 | -0.47 | 1.90 | 2.02 | 2.33 | 1.26 | 5.14 |
| Household head education — ref: No education |  |  |  |  |  |  |  |  |  |  |  |  |  |  |
| Primary | 0.02 | 0.10 | 0.04 | 0.15 | 0.16 | 0.14 | 0.20† | 0.11 | -0.04 | 0.10 | -0.11 | 0.09 | -0.07 | 0.07 |
| Secondary / higher | 0.11 | 0.15 | 0.31 | 0.24 | 0.10 | 0.20 | 0.20 | 0.19 | -0.04 | 0.16 | 0.10 | 0.13 | 0.08 | 0.11 |
| HH head female | -0.24* | 0.12 | -0.08 | 0.16 | -0.20* | 0.10 | -0.13 | 0.14 | -0.24* | 0.11 | -0.20* | 0.09 | -0.08 | 0.06 |
| HH members (n) | -0.04† | 0.02 | 0.02 | 0.04 | -0.07** | 0.03 | -0.02 | 0.03 | -0.03 | 0.02 | -0.07** | 0.02 | -0.06** | 0.02 |
| Residence — ref: Rural |  |  |  |  |  |  |  |  |  |  |  |  |  |  |
| Urban (ref: Rural) | 0.13 | 0.17 | 0.86** | 0.30 | 0.69** | 0.17 | 0.26 | 0.22 | -0.06 | 0.13 | 0.15 | 0.17 | 0.07 | 0.10 |

*\*\* p < 0.01 \* p < 0.05 † p < 0.10. RSE = robust standard error. Weighted quantile regression . Dependent variable: child haemoglobin (g/dL). OLS estimates from survey-weighted linear regression. Ethiopia DHS.*

**Table 4.**
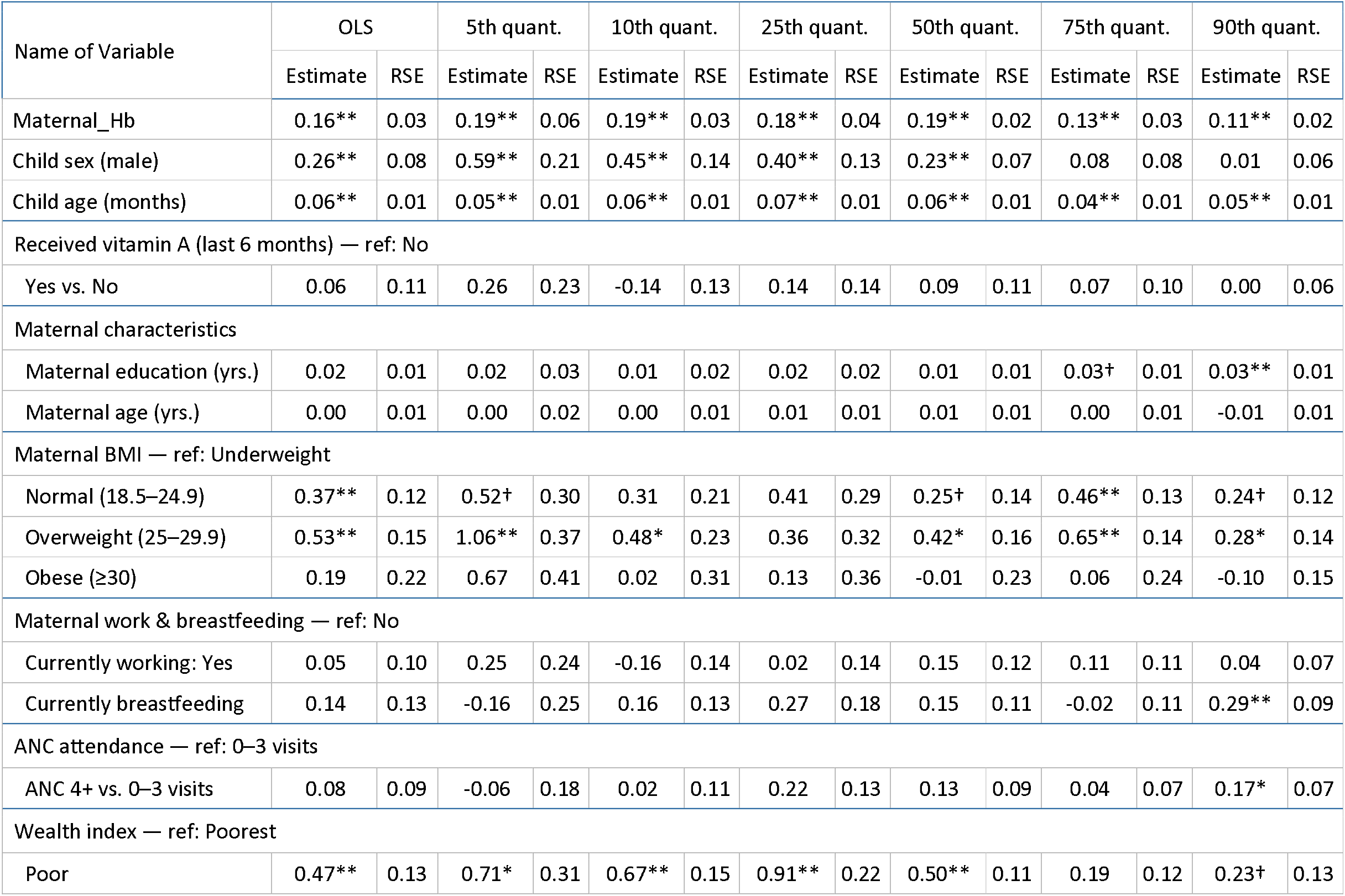
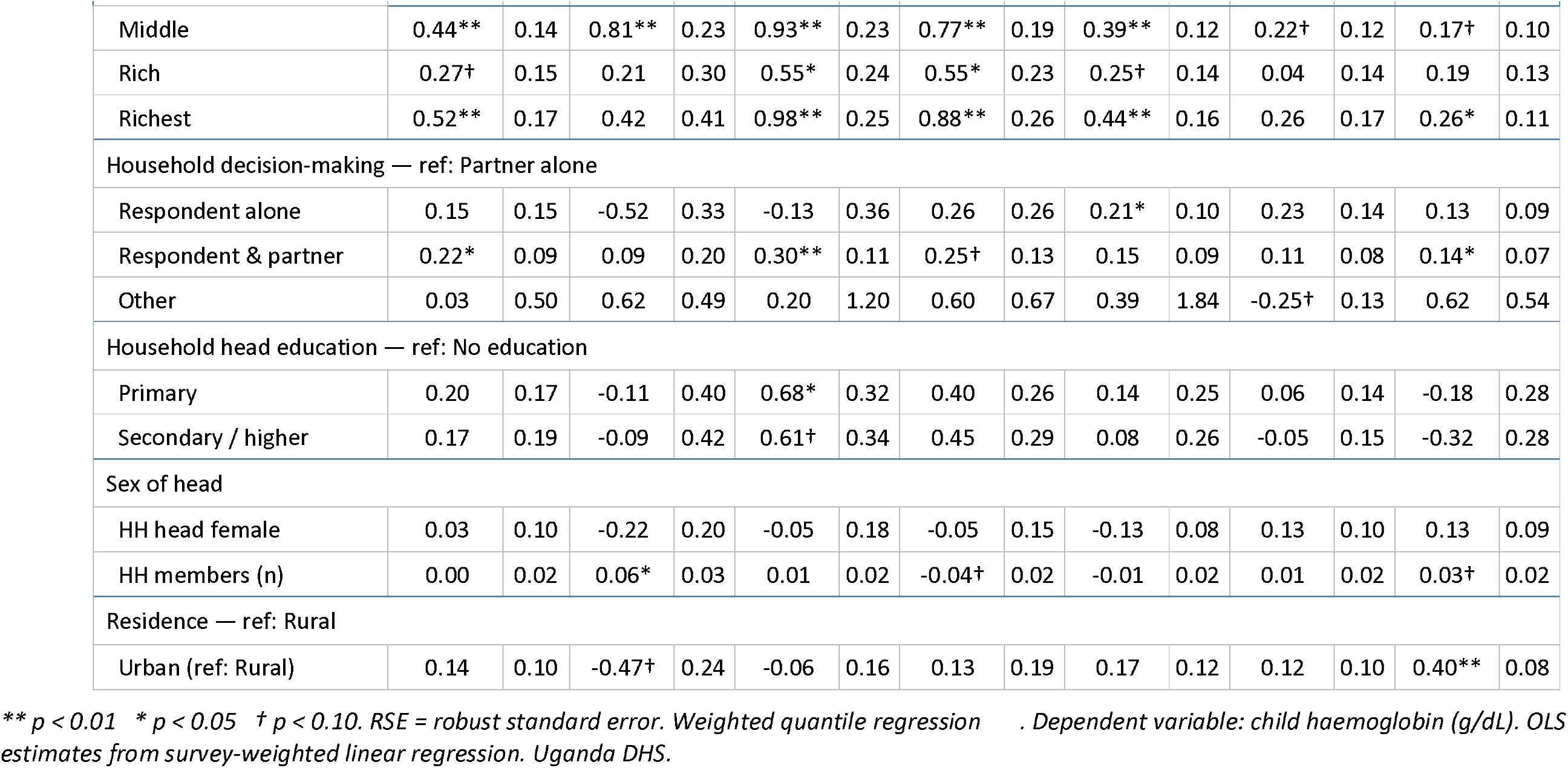
Multivariable analysis of the association between maternal and child haemoglobin concentration Uganda (N = 1,600).

### Graphical presentation of the results

Figures 1-3 present the results of the QR analysis conducted in the three countries. These graphs illustrate how maternal Hb concentration is associated with child Hb levels across different quantiles. Additionally, the graphs highlight that the magnitude of these effects at various quantiles differs significantly from the OLS coefficients, including the CIs around each coefficient.

**Figure 1:**
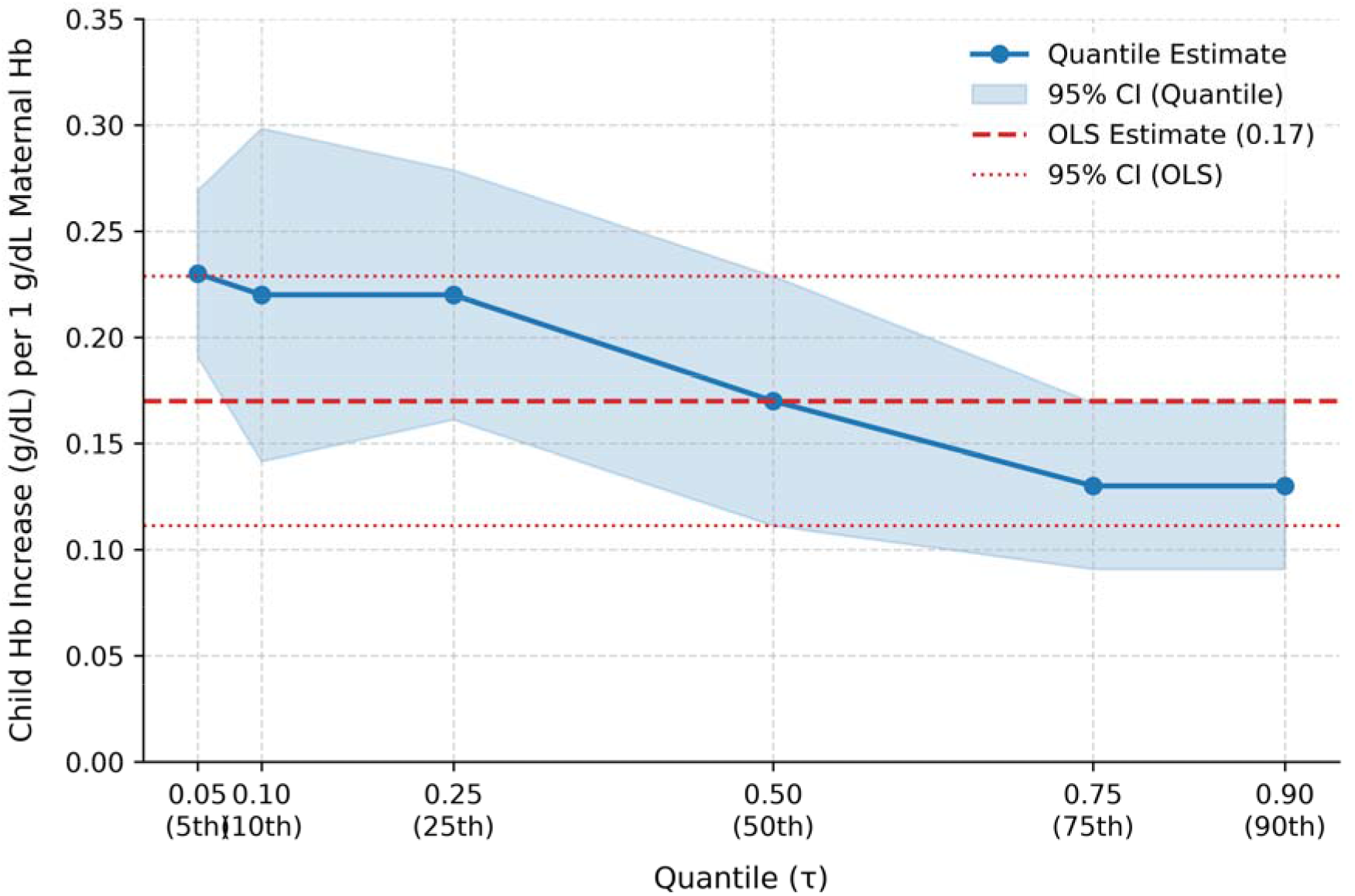
Association between maternal and child Hb concentration, Ethiopia.

**Figure 2:**
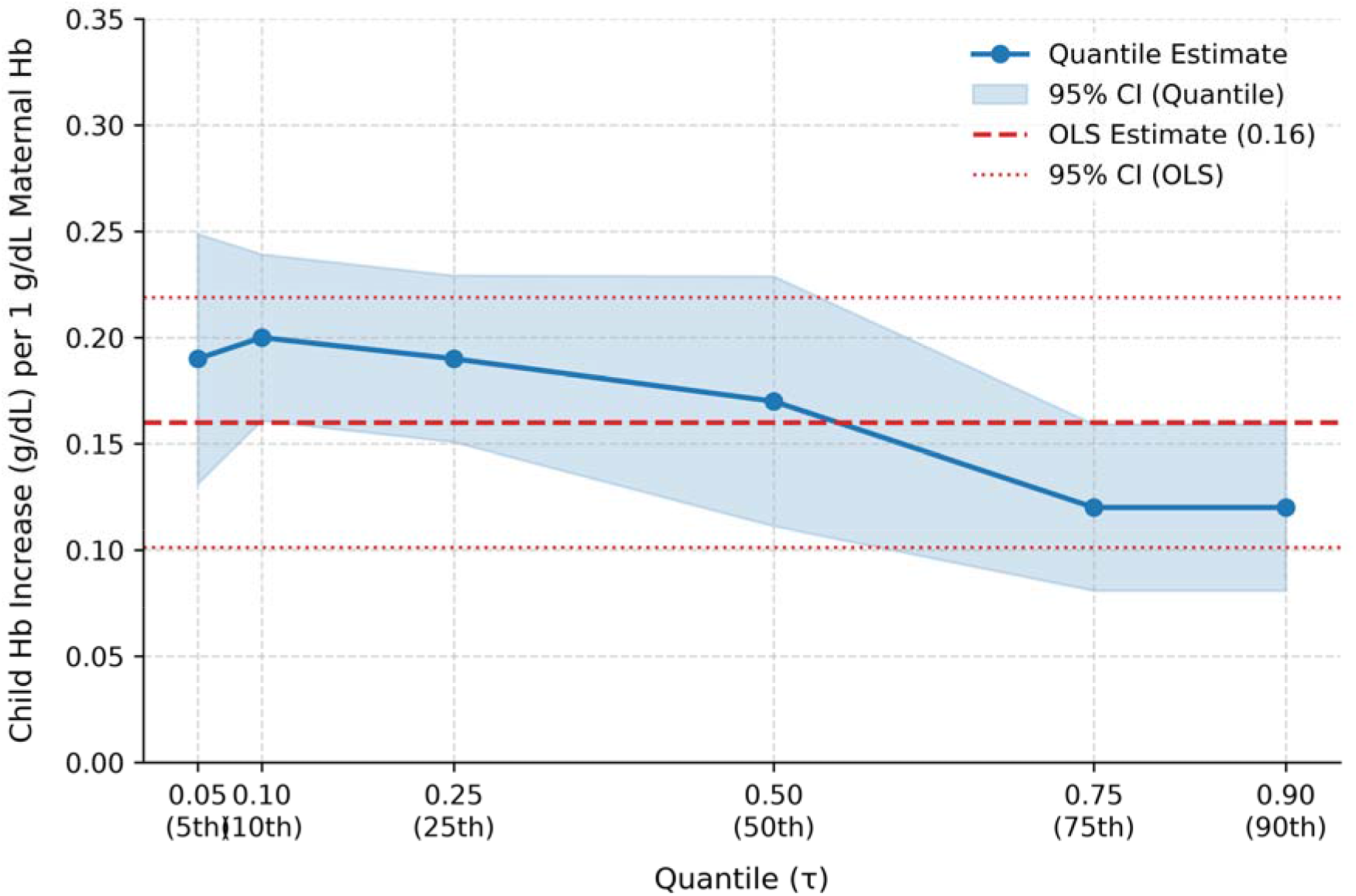
Association between maternal and child Hb concentration, Tanzania.

**Figure 3:**
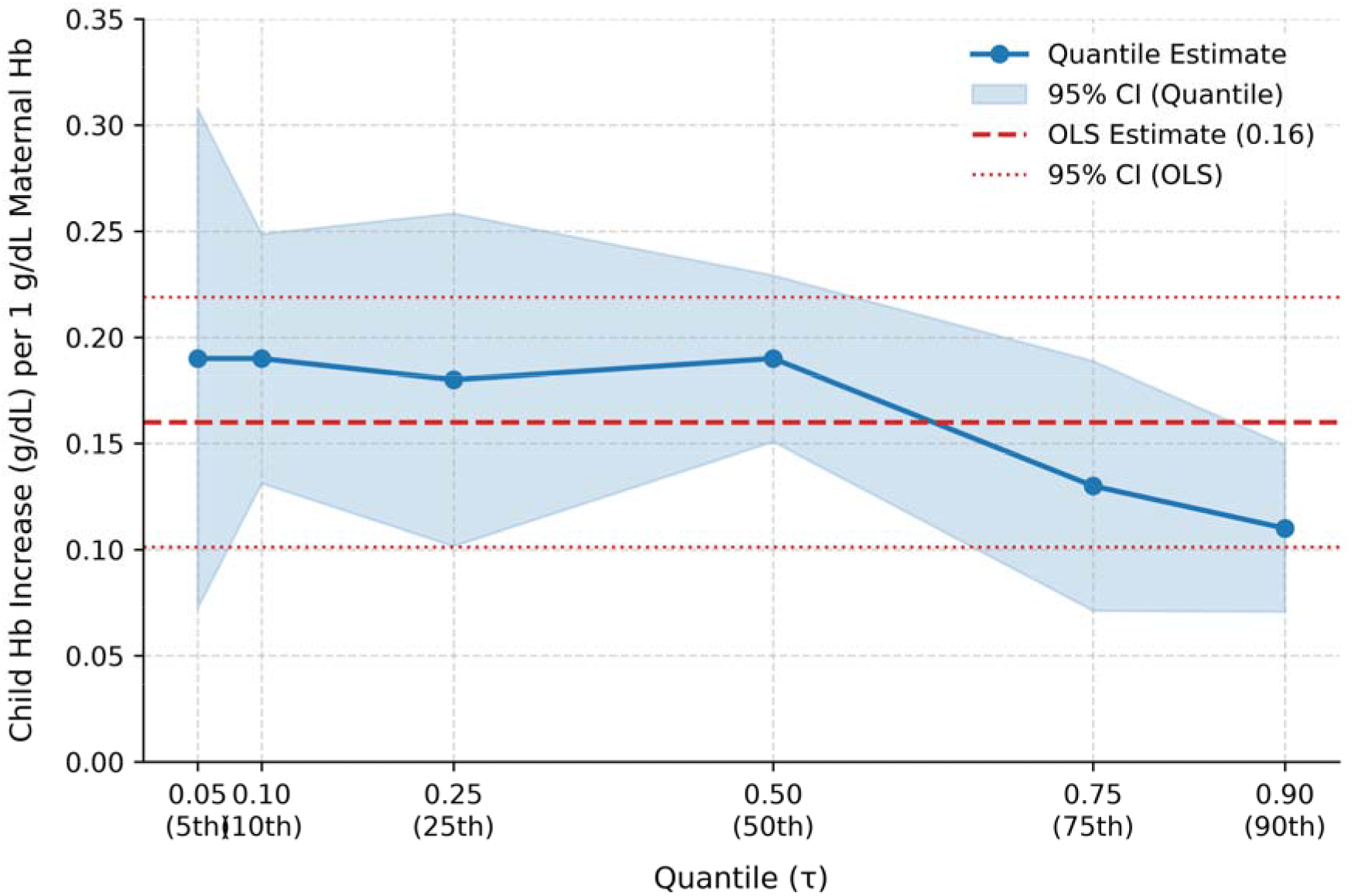
Association between maternal and child Hb concentration, Uganda.

## Discussion

This study investigates the relationship between maternal haemoglobin (Hb) concentration and child Hb levels in three Eastern African countries: Tanzania, Ethiopia, and Uganda. It uses quantile regression (QR) to analyse how the effect of maternal Hb on child Hb levels varies across the Hb distribution. Our findings reveal that maternal Hb concentration positively associated with children’s Hb levels across all quantiles. However, this effect diminishes as we move to higher quantiles. Notably, the strongest association is seen in children in the lower quantiles, who are at higher risk of anaemia, whereas the effect is minimal among those in the higher quantiles. These results suggest that improving maternal Hb concentration benefits children in lower quantiles more than those with higher quantiles. Therefore, interventions aimed at increasing maternal Hb concentration are likely to benefit children at the lower end of the Hb distribution more than those at the higher end.

The findings from QR analysis differ from those from OLS regression. While OLS analysis suggests a consistent positive mean association between maternal Hb concentration and children’s Hb levels across the three countries, QR findings reveal a more nuanced picture. These findings highlight the importance of accounting for the distributional effects of maternal Hb concentration, as relying solely on OLS can misrepresent its effects on child Hb levels. This research highlights a crucial link between maternal and child Hb concentration, underscoring the importance of addressing maternal anaemia to reduce children’s anaemia risk. By effectively improving the Hb concentration of expectant mothers and women of reproductive age, we can substantially reduce the risk of their children experiencing anaemia later in life. The findings discussed above can be attributed to several factors, particularly the socioeconomic and environmental conditions shared by children and their mothers. Several studies (17–25) show that these shared conditions significantly affect overall well-being and health.

The findings of this study are consistent with previous research on the effects of maternal Hb concentration on the Hb levels in children. Numerous studies conducted in low- and middle-income settings have documented these effects (3, 7–12, 17–20, 26). Specifically, children of non-anaemic mothers are less likely to be anaemic compared to those whose mothers are anaemic. One study found that only 1 in 4 children of non-anaemic mothers was anaemic, while the rate was 3 in 4 among children of anaemic mothers (3). Additionally, research has shown that children of mothers with high Hb concentration are less likely to experience anaemia compared to their peers with anaemic mothers (9). This direct connection between maternal anaemia and child anaemia highlights a cross-generational impact (8). The finding also reinforces the idea that children of non-anaemic mothers face a lower risk of developing elevated levels of anaemia compared to those whose mothers are anaemic (7–12). Consequently, improving mothers’ nutritional status is crucial to reducing high rates of child anaemia in low-income settings.

### Strengths and limitations

The study has several notable strengths. One key strength is that both the outcome variable (child Hb) and the candidate predictor variable (maternal Hb) were objectively measured, which helps minimise potential biases associated with subjective measurements. Using QR allowed assessment of the effects of maternal Hb concentration across the distribution of child Hb levels, providing a more comprehensive understanding of these effects. Additionally, using nationally representative data allows the results to be generalised to all children in the respective countries. However, an important limitation of the study is its cross-sectional nature, which makes it difficult to establish causality. Thus, the findings can only be interpreted as associations between the outcome and predictor variables. The term “effect” in the manuscript refers to a non-causal association, specifically how changes in the coefficient of the explanatory variable correspond to changes in the coefficient of the outcome variable. Lastly, missing data can often pose a significant challenge in secondary data analysis. Fortunately, due to the robustness of the DHS data management system, missing data was not an issue in our analysis.

### Conclusion

We used QR to examine the differential effects of maternal Hb concentration on children’s Hb levels while controlling for other covariates. Our analysis clearly demonstrated heterogamous association between maternal Hb concentration and child Hb levels at different points within the Hb distribution across all countries studied. The strongest association is observed among children at the lower quantiles who are at high risk of anaemia, whereas the effect is less pronounce among children at the higher quantiles. Therefore, improving maternal Hb is likely to benefit children in the lower quantiles more than those in the upper quantiles. Interventions and programmes aimed at addressing child anaemia should account for the varying effects of maternal Hb concentration on child Hb concentration across different Hb distribution quantiles.

## Acknowledgement

We wish to express our profound gratitude to The DHS Program, USA for providing us access to the data. We also wish to acknowledge institutions of respective countries that played critical roles in the data collection process.

## Data availability statement

The datasets generated and/or analysed during the current study are available in the DHS program repository, https://dhsprogram.com/data/available-datasets.cfm (accessed on 30/05/2026). Data are accessible free of charge upon a registration with the Demographic and Health Survey program (The DHS Program). The registration is done on the DHS website indicated above.

